# Chyme reinfusion using the Insides^®^ System to reduce parenteral nutrition dependence in Type 2 intestinal failure: multicentre randomised controlled trial (REINFUSE)

**DOI:** 10.64898/2026.04.26.26351226

**Authors:** T. Milne, S. Lal, A. Abraham, K. Farrer, J. Evans, K. Thomas, C. Vaizey, S. Gabe, D. Layfield, N. Randhawa, J. Rogers, D. Mercer, L. Barda, R. Carroll, M. Rosenthal, S. Jafri, C. Frampton, D. Burke

**Author notes:** **Corresponding Author** Tony GE Milne 5/31 Oakwal Terrace, Windsor 4030, Brisbane, QLD, Australia.

## Abstract

**Background:** Patients with type 2 intestinal failure (T2IF) due to double enterostomy or enterocutaneous fistula (DES/ECF) require parenteral nutrition (PN), carrying risks of catheter sepsis, venous thrombosis, and liver disease. Chyme reinfusion therapy (CRT) may reduce PN dependence but has not been evaluated in a randomised controlled trial (RCT). This study assessed whether device-assisted CRT using The Insides System reduces PN requirements.

**Methods:** This multicentre, open-label RCT enrolled PN-dependent adults with T2IF due to DES/ECF across 12 centres in the UK and USA. Patients with insufficient distal limb length, proximal bowel obstruction, active sepsis, or severe hepatorenal failure were excluded. Participants were block randomised 2:1 to device-assisted CRT plus standard care (active) or standard care (control). The primary endpoint was ≥50% reduction in PN caloric intake at 30 days, using an intention-to-treat analysis, for a comparison between randomised groups using a two tailed p-value of 0.025 to allow for a single interim analysis. Secondary outcomes were rate of PN cessation at 30 and 60 days, quality of life, and adverse events.

**Results:** The population comprised 39 (26 active, 13 control) participants. At Day 30, 8/26 (31%) active participants achieved the primary endpoint versus no controls (p=0.035). By Day 60, 10/23 (43%) active participants had completely ceased PN versus no controls (p=0.008), with median intestinal losses reduced by 1,344 mL/day at Day 30 (p=0.005) and 1,450 mL/day at Day 60 (p=0.026 between group). Device-related adverse events were predominantly mild; one death unrelated to the device occurred.

**Conclusion:** Although the study’s primary endpoint did not reach statistical significance, CRT with the Insides System demonstrated substantial therapeutic advantages in patients with T2IF from DES/ECF. At 30 days, 31% of participants had reduced PN calories by 50% and at 60 days >40% had completely ceased PN, with an acceptable safety profile.

**Trial registration:** ClinicalTrials.gov NCT04577456

**Funding:** This trial was sponsored by The Insides Company Ltd.

**Surgical Relevance:** *What is already known:* Patients with type 2 intestinal failure due to double enterostomy or enterocutaneous fistula depend on parenteral nutrition, which carries significant risks including central venous catheter (CVC) sepsis, venous thrombosis, and intestinal failure-associated liver disease. Chyme reinfusion therapy restores distal gut function but has only been evaluated in non-randomised cohort studies.

*What is new:* This first randomised controlled trial of device-assisted chyme reinfusion demonstrates that 43 per cent of participants can completely cease parenteral nutrition by 60 days, with a 70 per cent reduction in intestinal losses, high participant satisfaction and an acceptable risk profile.

*Potential impact on future practice:* Early initiation of device-assisted chyme reinfusion in suitable patients with double enterostomy or enterocutaneous fistula reduces parenteral nutrition dependence, avoids associated complications and costs, and facilitates rehabilitation before reconstructive surgery.

## Introduction

Intestinal failure, resulting from proximal double enterostomy or enterocutaneous fistula (DES/ECF), is a complex and life-threatening condition. Inadequate intestinal length leads to insufficient absorption of fluids and macronutrients to maintain normal homeostasis, with long-term parenteral nutrition (PN) requirement, and substantial associated morbidity (1,2). The rate of central venous catheter (CVC) related sepsis for patients on long-term PN ranges from 0.22-11.5 events per 1000 catheter days (3–5), with the presence of an ostomy and fistula potentially increasing the risk (6). Additional significant risks include central venous thrombosis (7), liver injury (8), and elevated mortality statistics of up to 39% at 10 years (9).

The sequelae of DES/ECF also extend to the healthcare system, with a profound economic burden. Home PN is expensive, costing up to GBP73,000 per year (10), while the total inpatient costs for type 2 intestinal failure (T2IF) on PN may reach GBP1000 per day (11). Furthermore, an episode of CVC-related sepsis may increase hospital stay, with additional unplanned admissions and complications, and complex stoma appliance maintenance potentially consuming significant further resources (12).

Chyme reinfusion therapy (CRT) involves refeeding proximal bowel enteric contents into the distal, or defunctioned, bowel, allowing functional restoration of distal gut activity for patients with DES/ECF. Although first applied systematically in the late 1970s (13), CRT did not achieve widespread use because of the difficulty of manual reinfusion of chyme and aversion to handling chyme (14). There have been sporadic case series of its use over the last 50 years, with the majority of this experience coming from France (15). These reports described the following potential advantages of CRT over standard PN based therapy for suitable patients with DES/ECF; cessation of PN in the majority of patients (14,15), reduced afferent stoma losses (16), improvement in liver dysfunction (15), restoration of nutritional autonomy (15,17), reduced morbidity after restorative surgery (18,19) and reduced healthcare spending (19). However, no randomised controlled trials (RCTs) have been performed to establish high-level evidence for the procedure.

In 2020, Sharma et al. reported a feasibility study of a novel chyme reinfusion device, The Insides^®^ System (The Insides Company, New Zealand), that provided a solution to the difficulties associated with manual chyme reinfusion and was safe, well-tolerated, and manageable for patients at home (20,21). Additional case series using the device have since been reported (22–24). If successful, this system would allow the routine reduction or cessation of home PN use for patients with DES/ECF, while potentially reducing healthcare spending. This multicentre RCT aimed to determine whether distal chyme reinfusion using The Insides System reduces the need for PN in patients with T2IF due to a proximal DES/ECF.

## Methods

This was a multicentre, multinational RCT reporting in accordance with the CONSORT 2025 statement (Supplementary Table 1). Participants were enrolled between October 2022 and August 2025 from 12 participating centres in the UK and the USA (Supplementary Table 2). Ethical approval was obtained for all participating study centres, and the trial was conducted in accordance with the Helsinki Declaration. All participants provided written informed consent. The trial protocol was prospectively registered with ClinicalTrials.gov (NCT04577456) before commencement of participant enrolment.

### Inclusion and exclusion criteria

To be eligible for inclusion, participants with DES/ECF had to be: aged 21 years and older, able to provide written consent, have exposed proximal and distal DES/ECF limbs on their abdominal wall, be dependent on PN, have had their DES/ECF for at least 2 weeks, and have a distal limb of their DES/ECF that can be intubated with a 24Fr feeding tube. Participants were excluded if they had insufficient access to the distal limb, proximal bowel obstruction, evidence of active *Clostridium difficile* infection or small bowel bacterial overgrowth, or signs of active systemic infection. Additional exclusion criteria were pre-existing gastrointestinal motility disorders (functional or metabolic), formation of an ileal pouch, known peritoneal malignancy, liver cirrhosis, coagulopathy, severe chronic kidney disease, an active implanted cardiac or neuromodulatory medical device, and patients who were pregnant or breastfeeding.

### Participant recruitment and assessment

Demographic information was collected, including age, sex, ethnicity (approved only in US), medical and surgical history, oral diet and PN regimens, and medications. All participants underwent a comprehensive assessment of their nutritional status, including vital signs, BMI, and serum blood tests, as well as evaluation for *C. difficile* infection. Eligible participants also underwent a contrast study to demonstrate distal bowel integrity and patency before enrolment. Enrolled participants also provided baseline data on their healthcare utilisation and quality of life using the EuroQol 5D-3L, Stoma-QoL, and Beck Depression Inventory questionnaires, as well as baseline handgrip strength.

### Randomisation and blinding

Eligible participants were randomised centrally, using a computer generated randomisation sequence, in a 2:1 ratio to receive either chyme reinfusion using The Insides System plus standard care (active group) or standard (PN) therapy (control group) as per the ASPEN and ESPEN treatment guidelines (25,26). Participants were randomised in blocks of 6 and stratified based on whether they had ECF or DES (± ECF). Due to the nature of this study, the participants and the treating clinicians were not blinded to the intervention.

### Study outcomes and data collection

The primary endpoint was the proportion of participants achieving a 50% reduction in caloric intake from PN by 30 days post randomisation in a modified intention-to-treat (mITT) analysis. MITT excluded patients who were unable to have a feeding tube inserted into the distal enterostomy, however no ITT participants were excluded from the mITT population. Consequently, mITT and ITT results are identical and are reported as ITT. PN requirements were captured for the 7 days before randomisation and were assessed daily for up to 60 days post-randomisation for assessment of the 7 days periods comprising the primary and secondary endpoints.

Secondary outcomes included time to reduction or discontinuation of PN, length of hospital stay, quality of life measures, satisfaction scores, serum blood tests, Nutrition Risk Index (NRI) (27), date of DES/ECF closure, chyme losses, and incidence of adverse events. The NRI was calculated using the following formula: NRI = (1.519 × albumin g/dL) + (41.7 × current weight / usual weight). A score >100 indicates no risk, 97.5–100 mild risk, 83.5–97.4 moderate risk, and <83.5 severe risk. Secondary endpoints were recorded at 30 days and 60 days post-randomisation. Feedback on the use of the device was assessed using a custom-designed satisfaction questionnaire (Supplementary Table 3). Adverse events were recorded using the CTCAE v4.03 classification system (28) and were monitored by an independent Data Safety Monitoring Board (DSMB). Participant withdrawals and protocol deviations were recorded. Data were entered directly into the TrialPoint™ electronic data capture system hosted by Databean, Inc. (St. Augustine, FL, USA) and independently monitored for compliance with the study protocol and accuracy with source documentation.

Participants allocated to the active group had the placement of a distal limb chyme reinfusion tube following randomisation. All participants were instructed on how to use The Insides System, which consists of a custom 22Fr or 28Fr distal catheter, a small impeller pump that connects to the catheter within the stoma appliance, and an external battery-operated driver that couples with the impeller to drive the pump (Figure 1). Device safety and feasibility, and reinfusion bacterial load safety have all been previously established for this system (20). The pump was changed every 3 days, and the reinfusion tube was changed every 30 days by a healthcare professional.

**Figure 1.**
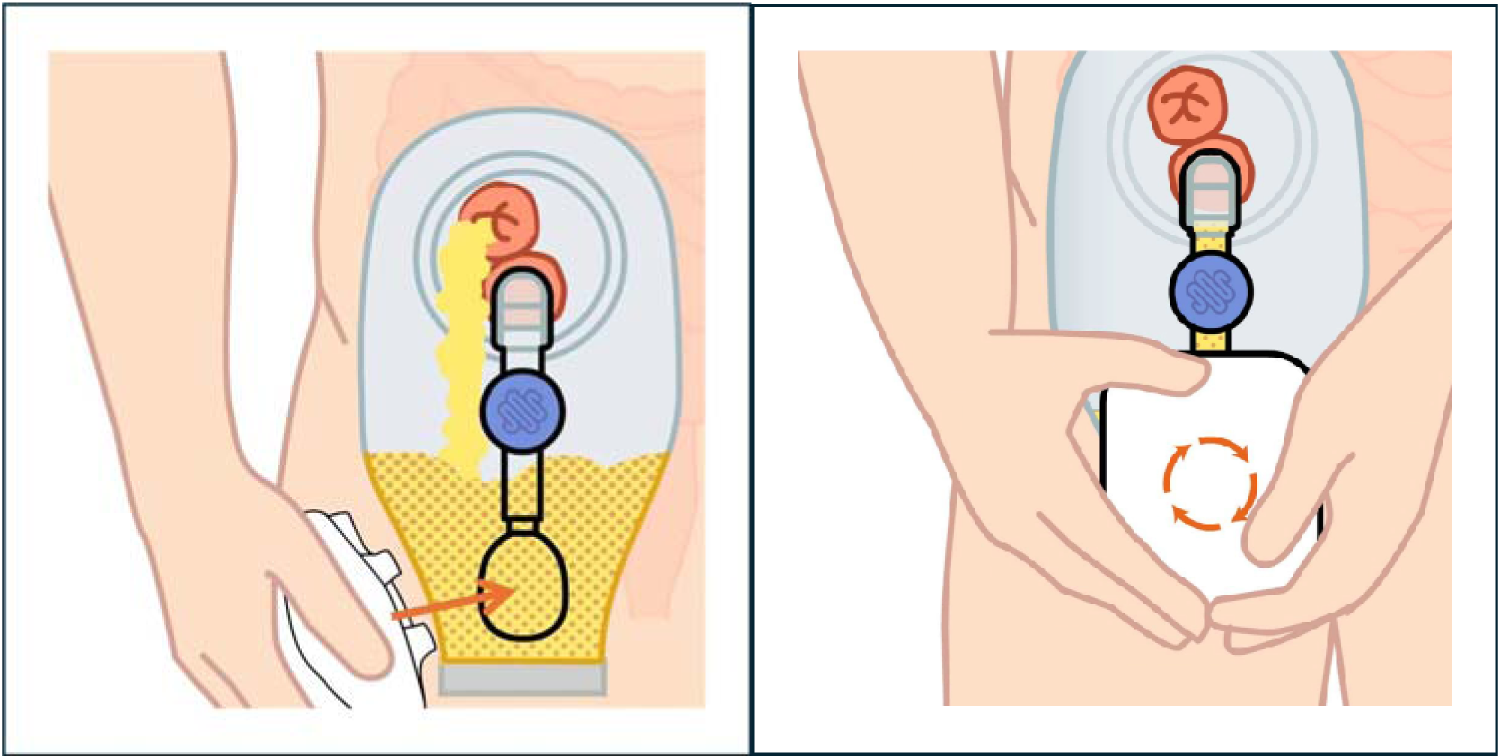
Schematic illustration of the method of use of the Insides System device.

During the 60-day study period post-randomisation, PN and other intravenous therapy use, chyme and stool production, medication use, and diet were recorded daily. All participants were assessed weekly to determine eligibility for weaning from PN to oral/enteral nutrition (EN) according to the guidelines summarised in Supplementary Figure 1. Formal assessments occurred at 30 days and 60 days post randomisation and prior to DES/ECF closure to capture NRI score (27), a complete nutritional evaluation including biochemical markers, quality of life assessments, grip strength, and utilisation of healthcare services. Adverse events were recorded daily with oversight by the independent medical monitor and DSMB. After the conclusion of the 60-day follow-up study period, participants from both groups were given the option to use the refeeding system until DES/ECF closure, as permitted by applicable regulatory approvals.

### Sample size calculation and statistical analysis

The following estimates were used to calculate the sample size for this study, 10% of control participants and 65% of the active group would achieve the primary endpoint (17,20). Using power of 90%, 39 participants were required at a 2:1 allocation ratio to show this difference as statistically significant with a two-tailed α=0.05. An interim analysis of the primary endpoint was performed to assess whether efficacy had been achieved early. The interim analysis and the final analysis were each conducted at the 0.025 level (2-sided), using the Bonferroni adjustment, thereby maintaining an overall Type I error rate of 0.05.

The primary analyses were undertaken using the mITT/ITT population. An as-treated (AT) subgroup analysis was performed for safety endpoints. Statistics were conducted using SPSS v30.0. The analyses of the proportion of participants achieving a 50% reduction in caloric intake from PN were undertaken using a 2-sided Fisher’s Exact Test. Comparisons of continuous outcome measures between randomised groups were made using Mann-Whitney U tests and Kaplan Meier curves and log-rank tests were used to summarise and compare the time to cessation of PN. All p-values presented for secondary endpoints are nominal.

## Results

Forty-four participants were enrolled, of whom 39 participants were randomised (26 active and 13 control), as reported in the CONSORT diagram (Figure 2). Three participants in the active group did not have Day 30 PN data recorded (one died of underlying complications, one was unable to maintain bag adherence, and one was unable to retain the feeding tube), but were retained in the mITT analysis, giving a mITT/ITT population of 39 (26 active, 13 control). Two control participants were withdrawn between Day 30 and Day 60, one by participant choice, and one following transition to palliative care. Baseline characteristics of the two groups are reported in Table 1. Mean age was 57.5 years and 49% were female. The median length of the proximal small intestine was 77.5 [interquartile range (IQR) 40-147.5] cm, and of the distal small intestine was 123 [70-180] cm. Of the 39 participants, 29 had some residual colon. All but 2 randomised participants were recruited by the UK trial centres (Supplementary Table 2).

**Figure 2.**
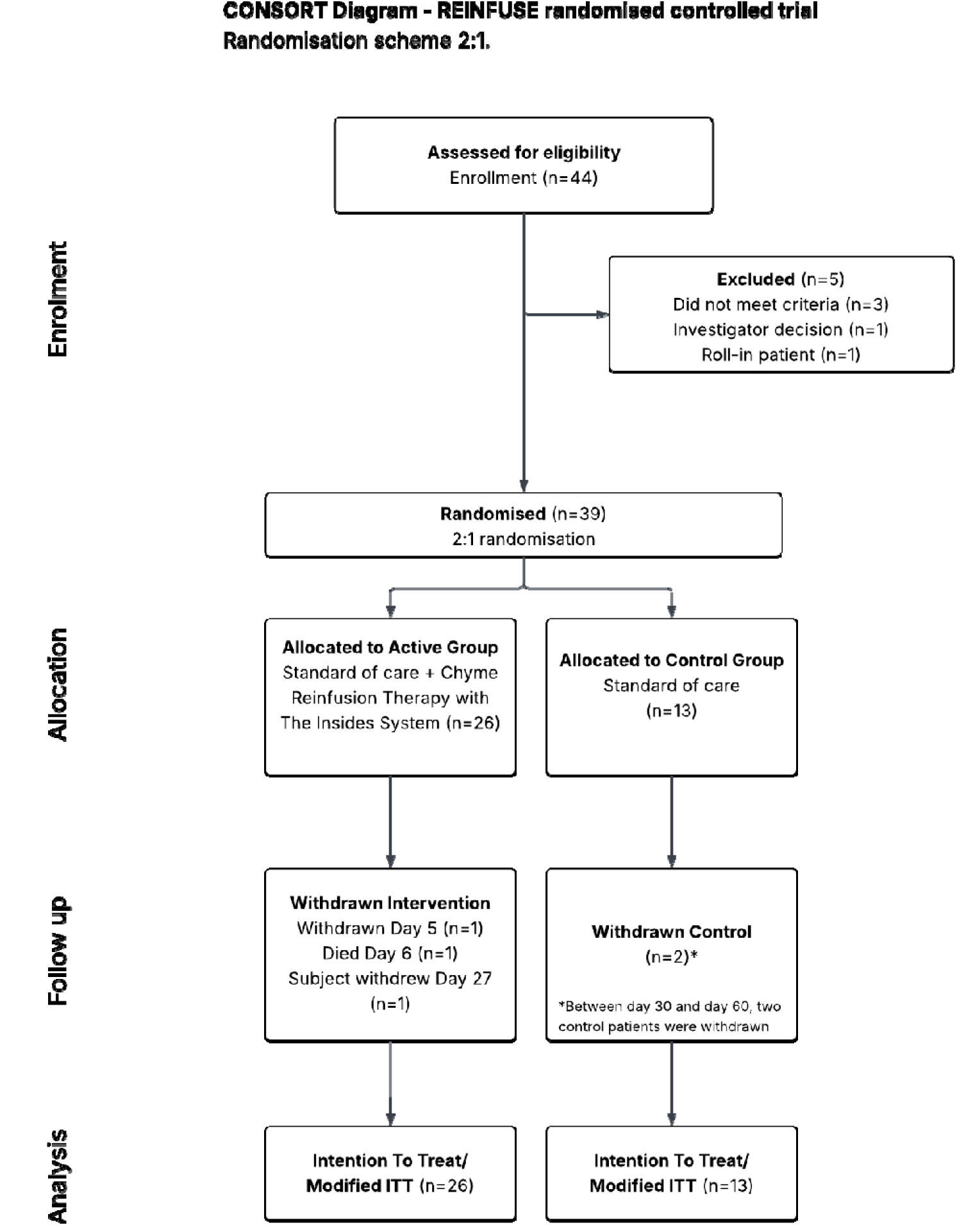
CONSORT Diagram for the REINFUSE trial.

**Table 1.** Baseline Characteristics.

|  | **Active Group**<br>**(N=26)** | **Control Group**<br>**(N=13)** | **Total (N=39)** |
| --- | --- | --- | --- |
| **Age (years)** |  |  |  |
| Mean (SD) | 55.1 (13.9) | 62.2 (11.2) | 57.5 (13.3) |
| **Gender N (%)** |  |  |  |
| Female | 13 (50) | 6 (46.2) | 19 (48.7) |
| Male | 13 (50) | 7 (53.8) | 20 (51.3) |
| **BMI (kg/m<sup>2</sup>)** |  |  |  |
| Mean (SD) | 24.7 (5.1) | 23.9 (4.2) | 24.4 (4.8) |
| **Medical conditions N (%)** |  |  |  |
| Yes | 22 (84.6) | 12 (92.3) | 34 (87.2) |
| **Major comorbidities N (%)** |  |  |  |
| Malignancy of abdominal organ | 3 (11.5) | 1 (7.7) | 4 (10.3) |
| Crohn's disease | 8 (30.8) | 2 (15.4) | 10 (25.6) |
| Inflammatory bowel disease | 2 (7.7) | 0 (0.0) | 2 (5.1) |
| Other autoimmune | 4 (15.4) | 2 (15.4) | 6 (15.4) |
| Other abdominal disease | 8 (30.8) | 8 (61.5) | 16 (41) |
| **Hospitalised at enrolment N (%)** |  |  |  |
| Yes | 12 (46.2) | 8 (61.5) | 20 (51.3) |
| **Ostomy/fistula N (%)** |  |  |  |
| DES | 23 (88.5) | 10 (76.9) | 33 (84.6) |
| ECF | 3 (11.5) | 2 (15.4) | 5 (12.8) |
| Both DES/ECF | 0 (0) | 1 (7.7) | 1 (2.5) |
| **Proximal small bowel length (cm)** |  |  |  |
| Median [LQ, UQ] | 70 [40, 120] | 80 [35, 150] | 77.5 [40, 147.5] |
| **Distal small bowel length (cm)** |  |  |  |
| Median [LQ, UQ] | 124 [82, 182.5] | 120 [64, 130] | 123 [70, 180] |
| **Colon N (%)** |  |  |  |
| Yes | 17 (65.4) | 12 (92.3) | 29 (74.4) |
| No | 9 (34.6) | 1 (7.7) | 10 (25.6) |
| **Baseline chyme output** |  |  |  |
| Median [LQ, UQ] | 2215.0 [1178.3, 2960.7] | 1468.2 [1072.5, 2801.0] | 1963.6 [1132.2, 2884.3] |

Chyme reinfusion was successfully continued to Day 30 or death in 23 (88%) active group participants. At 30 days, eight (31%) of the 26 active group participants had their PN calorie requirement reduced by ≥50%, compared to none (0%) of the 13 participants in the control group (p=0.035). This improvement did not meet the pre-specified trial endpoint based on the interim analysis adjustment to p<0.025. Although the protocol assumed that patients in the active group began CRT on the day of randomisation this was not the case for several participants. A sensitivity analysis comparing the percentage reaching a 50% reduction in PN at day 30 following the start of therapy, indicated a significant difference between active and control groups (9/26 v 0/13, p=0.0175). By Day 60, the difference between groups was more pronounced; 10 (38%) of the 26 active participants had PN calorie prescription reduced by ≥50%, compared to none (0%) in the control group (38% vs 0% p=0.016), with 10 (Kaplan-Meier estimate=43%) completely weaned off PN versus 0% of the control group (log-rank test p=0.008) (Figure 3).

**Figure 3.**
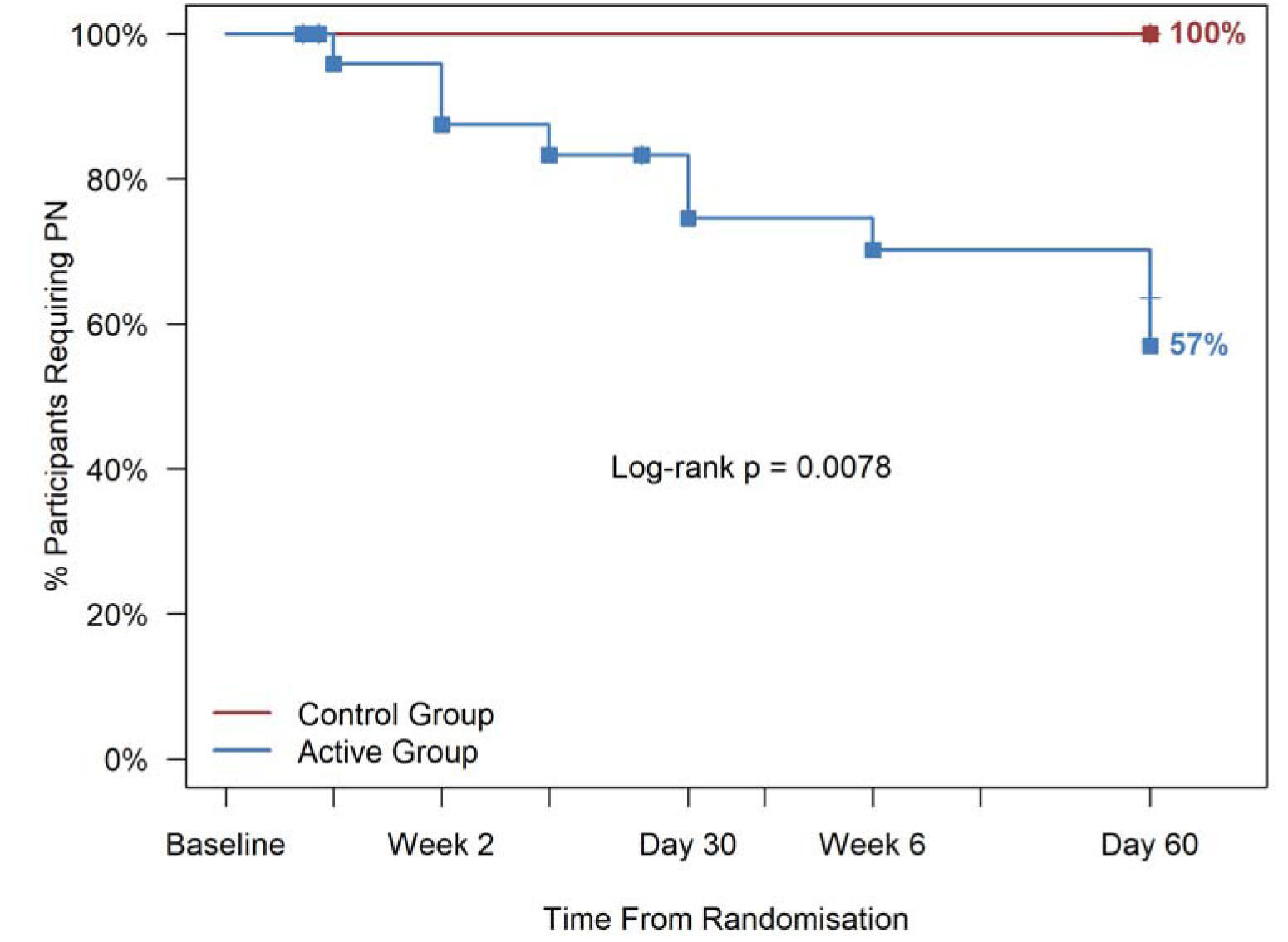
Kaplan Meier plot showing cumulative percent of participants completely weaned off parenteral nutrition at 60 days of therapy (log-rank p=0.008).

Device-assisted CRT was associated with a substantial and sustained reduction in intestinal losses in the active group over the 60-day study period (Figure 4). Change from baseline was analysed for participants with data at both timepoints. By Day 30, the active group had a median reduction of -1,344.3 mL/day [IQR: -2414.3, -265.7] (n=19), compared to -33.9 mL/day in controls [IQR: -600.0, 88.6] (n=11) (Mann-Whitney p=0.005). This treatment effect was maintained through Day 60, with a median reduction of -1,450.0 mL/day [IQR: -2439.1, - 458.6] (n=17) in the active group versus -157.3 mL/day [IQR: -1083.3, 60.4] (n=8) in controls (p=0.026).

**Figure 4.**
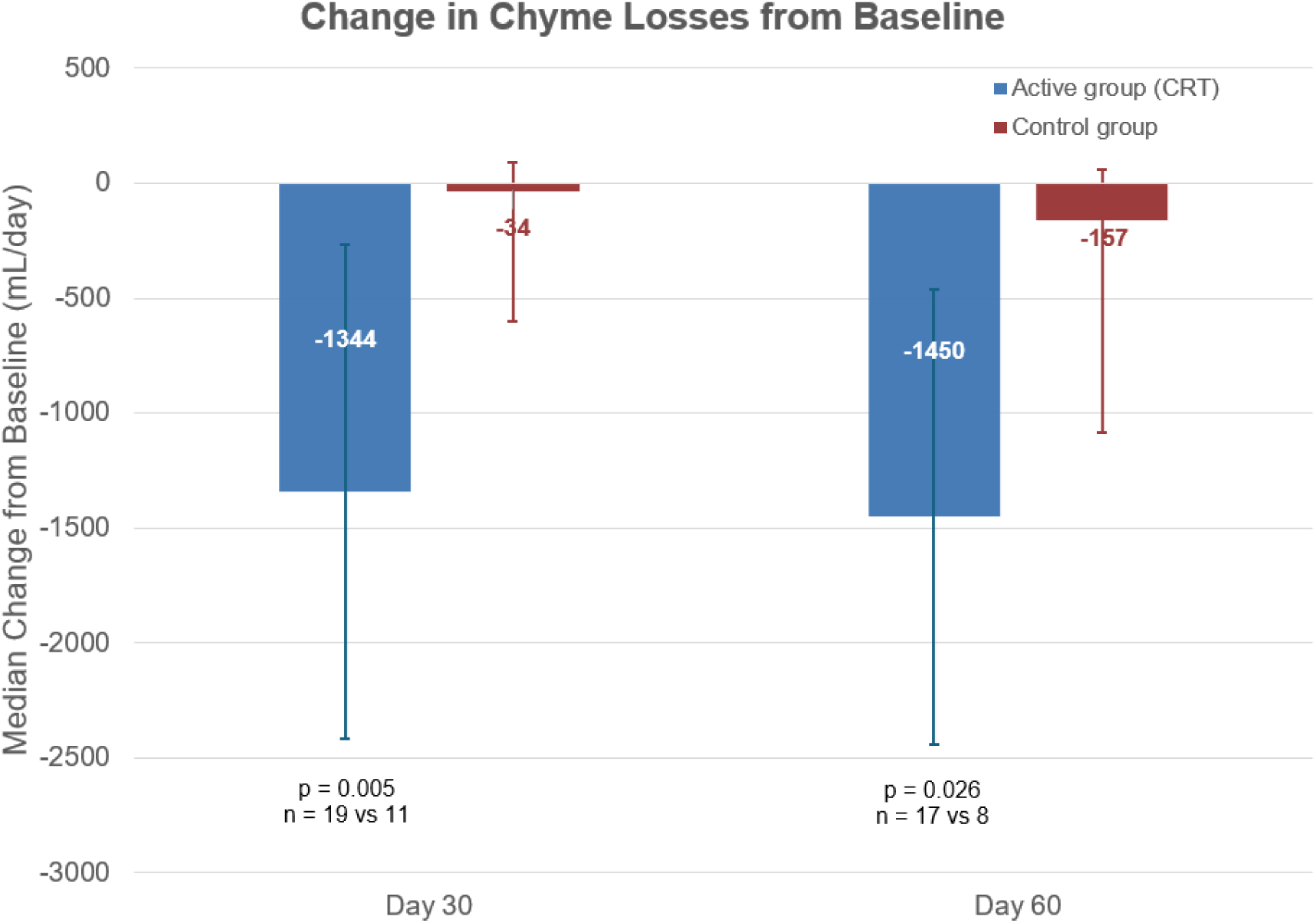
Median change in chyme losses from baseline (mL/day) with interquartile range (error bars) for active (CRT) and control groups. Analysis includes only participants with paired baseline and follow-up data. Day 30: active n=19, control n=11; Day 60: active n=17, control n=8. Participants with missing baseline data, withdrawal or death were excluded (active n=7, control n=1). Additional exclusions at Day 60 due to stoma closure, withdrawal, or incomplete data collection (active n=1, control n=4). p-values from Mann-Whitney U test.

Results of liver enzymes and inflammatory markers are shown in Supplementary Figure 2. There were no notable changes between groups for the liver enzymes and inflammatory markers.

Figure 5 demonstrates the median NRI trajectories of the two groups. Median NRI of the active group increased from 97.3 (IQR 93.3, 112.3) at baseline to 107.3 (IQR 96.1, 113.0) at Day 60. The control group median was 102.7 (IQR 93.3, 105.9) at baseline and 98 (IQR 94.6, 109.9) at Day 60 (p=0.173, between group comparison of the changes). The proportion of participants classified as no malnutrition risk increased from 43.5% to 66.7% in the active group, while decreasing from 53.8% to 40.0% in the control group.

**Figure 5.**
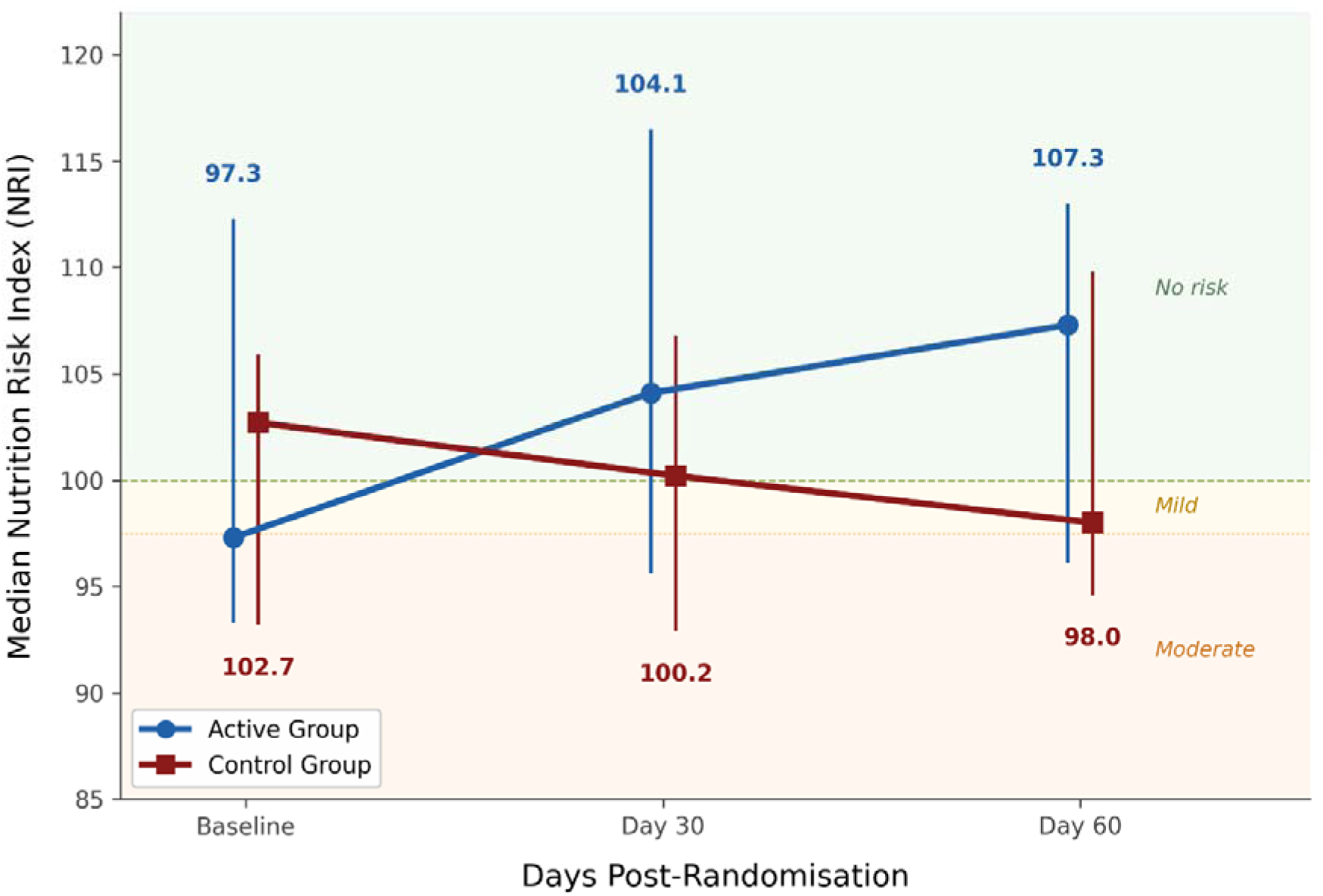
Median Nutrition Risk Index (NRI) with interquartile range (error bars) at Baseline, Day 30, and Day 60 for active (CRT) and control groups. Background shading indicates NRI risk categories: no risk (>100), mild risk (97.5–100), moderate risk (<97.5). Trends between groups remained similar (Mann-Whitney U p=0.173).

Of 39 participants, twenty were hospitalised at enrolment (12 active and 8 control). The median length of hospital stay was similar between groups (active 16.1 days vs. control 16.0 days).

### Device Satisfaction and Quality of Life

Participants in the active group reported very high satisfaction with using the Insides System (median 46/50 in a customised 10 question device evaluation survey, measured at Day 60 – (Supplementary Table 4). EuroQol-5D-3L and Stoma QOL Scale scores, and Beck Depression Inventory scores were comparable between the groups and over time (Supplementary Table 5).

### Safety Data

Safety analyses are reported in the AT population which, unlike all other results, included a non-randomised active roll-in participant (Figure 1). Between consent and Day 60, non-device-related adverse events were similar in frequency between groups: 24 events in 19 participants (70.4%) in the active group compared with 15 events in six participants (46.2%) in the control group (Supplementary Table 6, Supplementary Table 7). Total serious adverse events (SAEs) were more frequent in the active group with 14 events (2 device related) in 8 participants versus 2 events in 1 control participant. One participant in the active group died on Day 5 from mesenteric and hepatic vascular compromise secondary to occlusion of pre-existing coeliac and superior mesenteric artery stents. The event was reviewed by the DSMB and adjudicated by the independent medical monitor as unrelated to the device or study procedure.

By Day 60, 11 device-related adverse events were recorded in 9 of 27 active group participants; six were mild, four moderate, and one severe (refer Supplementary Table 8). Two device-related events were classified as SAEs, both involving peristomal skin irritation that required hospitalisation. The most common device-related events were reinfusion site conditions related to the feeding tube (five events in three participants) and abdominal pain (one participant experienced discomfort while pumping which resolved within 3 weeks, another found the in-situ tube painful and requested it removed). At 60 days two participants still had skin erosions, but all other device-related events had resolved. No device-related events required surgical intervention during this period and no unanticipated adverse device effects were reported.

## Discussion

This multicentre RCT did not meet the pre-specified trial endpoint of 50% reduction of PN calories at 30 days at p<0.025. However, The Insides System did enable 43% of participants with T2IF due to DES/ECF to completely cease PN within 60 days, compared with none in the control group. Median stoma/fistula losses also reduced by 70% from baseline in the active group, with no change in the control group. Device-related adverse events were predominantly mild, with only two instances of peristomal skin irritation unresolved at Day 60. Quality of life and depression scores were comparable, and participants utilising the Insides device expressed a high level of satisfaction in the technique and confidence in independent use.

These results have substantial clinical implications for the management of T2IF due to DES/ECF. Cessation or reduction of prolonged PN directly mitigates many of its well-documented consequences; including CVC-related sepsis, central vein thrombosis, and intestinal failure-associated liver disease (IFALD) (17). Beyond PN avoidance, CRT has been reported to have substantial additional benefits. It maintains trophic stimulation of the distal bowel, with a recent meta-analysis showing reduction in overall complications and improved gut recovery following reconstructive surgery, indicating that early device-assisted CRT can optimise patients for definitive restoration of continuity (19,29). Allowing the restoration of normal oral intake may provide advantages for psychological wellbeing and social participation (17). Substantial cost saving are likely, relating to reduced requirements of PN and home PN (HPN) services, and less time in hospital as patients provide their own nutrition at home while awaiting reconstruction (30,31).

The present study rate of cessation of PN of 43% at 60 days is lower than 85% cessation that was seen in five of the six cohort studies from a recent meta-analysis (14). Recognising that it is difficult to draw conclusions between studies where patient characteristics, underlying diseases and treatment protocols are not matched, there are two possible reasons for this difference. First, Picot et al, the authors of the largest study from France, have extensive experience in weaning patients off PN and are confident in doing so quickly. Sites included in the present study were mostly new to CRT and PN reductions only occurred once weekly (15). Second, patients in the cohorts that weaned with higher rates had much shorter intervals from initial surgery to starting CRT (Picot 33 days vs 92 days in the current trial’s active group) (17). This is supported by the fact that a further study by Farrer et al, including 22 patients with a median period of over 600 days from surgery to CRT, achieved cessation of PN in only 2 patients (24). These findings suggest that CRT should be initiated early in the patient journey to maximize weaning before the longstanding effects of diversion on the distal small bowel and colon are experienced.

While CRT is recommended in treatment guidelines (ASPEN, ESPEN) it has not yet gained wide utilisation because it is labour intensive and unpleasant for patients and their carers (29,32,33). The present study has demonstrated a high patient satisfaction rate for the use of The Insides System to perform CRT. This novel approach offers clinicians a more simple and acceptable method for introducing CRT to appropriate patients.

Reducing intestinal losses is critical in managing intestinal failure patients with DES and ECF (25,26). The study demonstrated a dramatic reduction in intestinal losses in the active participants compared to the controls at both 30 and 60 days. This is due to both the absorption of nutrients by the distal bowel and re-engagement of the ileal brake (13). When intestinal continuity is restored (via closure surgery or initiation of chyme reinfusion), enteroendocrine hormones Peptide YY (PYY) and GLP-1 are re-activated in the terminal ileum with chyme contact (34). This slows gastric secretions, increasing nutrient absorption within the small intestine and slowing the output from the DES and ECF. Achieving a reduction in intestinal losses also assists patient management, improving stoma bag adherence, and improving quality of life for the patient (35).

While the number of SAEs in the active group was higher than that in the control group, only two of these were related to the device, both involving peristomal skin excoriation. This imbalance should be interpreted in the context of care setting, as a lower proportion of active group participants were hospitalised at the time of AE onset compared with the control group. Active group participants managed in the community required hospital admission for events that would have been managed within existing inpatient care for hospitalised controls, inflating the active group SAE count. Four active group participants accounted for ten of the 14 SAEs.

The major strengths of this study are its multicentre nature, the low rate of withdrawals and complete data capture. It is the first RCT performed in this relatively rare condition and represents the strongest evidence to date that device-assisted CRT allows cessation of PN for many patients with high output DES/ECF. However, some limitations deserve mention. Despite the fact that treatment guidelines for T2IF are comparable between USA and UK/EU (25,26), only two USA participants were randomised from five sites. This may be due to the lack of previous experience of CRT nationally. Nevertheless, it is anticipated that the benefits demonstrated here will scale internationally, including into the US healthcare system. A further limitation was that the study’s primary endpoint did not reach statistical significance. There are several important reasons for this. First, the performance of a midpoint interim analysis required an adjustment of the p-value to <0.025. Second, several active patients did not begin CRT on the day of randomisation. To explore the impact of this, a sensitivity analysis comparing the percentage reaching a 50% reduction in PN at day 30 following the start of therapy was performed. This demonstrated a significant difference between active and control groups (9/26 v 0/13, p=0.0175). Third, the primary endpoint timing was informed by results from the largest cohort studies in France, where complete weaning was achieved in >85% of patients and usually before seven days (15,17). However, as mentioned above, the UK trial centres enrolled participants who had been on PN for more prolonged periods, and were slower to adjust PN doses than in historic French centres already familiar with CRT. Nevertheless, the compelling trial outcome data at both 30 and 60 days demonstrate important advantages for chyme reinfusion and PN reduction in these complex patients.

Future research should explore the broader therapeutic applications of device-assisted CRT beyond reduction of PN requirements. The marked reduction in intestinal losses observed in this study indicate that trials assessing the efficacy of CRT in reducing or eliminating intravenous fluid dependence in patients with high-output DES/ECF also deserve implementation. The role of device-assisted CRT as a rehabilitative tool to maintain or restore distal bowel function prior to reconstructive surgery requires further investigation, preferably in an RCT. In addition, studies that assess whether preoperative device-assisted CRT can predict post-operative anorectal function should be performed. These could offer clinicians and patients vital information when making decisions about stoma closure.

In conclusion, this first RCT of device-assisted CRT demonstrates safety and efficacy of a new method to achieve complete weaning from PN in many patients with T2IF secondary to DES and/or ECF. Device-assisted CRT also greatly reduces intestinal losses, is well tolerated by patients, with high satisfaction, and may improve nutritional status while offering strong risk-benefit justification. Application of this new approach in appropriate patients has the potential to become standard of care by aiding in the avoidance of PN related complications, rehabilitation of the distal bowel before restorative surgery, improving patient psychology, and ultimately reducing cost and hospital stay for these serious conditions.

## Data Availability

All data produced in the present study are available upon reasonable request to the authors

## Category

Randomised clinical trial

## Disclosure

The authors declare no conflict of interest.

This paper has been submitted for oral presentations at the ACPGBI meeting 2026 in Belfast, Ireland, and the Colorectal Spring Meeting 2026 in Cairns, Australia.

