## Supplementary Tables and Figures for "Chyme reinfusion using the Insides^®^ System to reduce parenteral nutrition dependence in Type 2 intestinal failure: multicentre randomised controlled trial (REINFUSE)"

**Supplementary Figure 1**

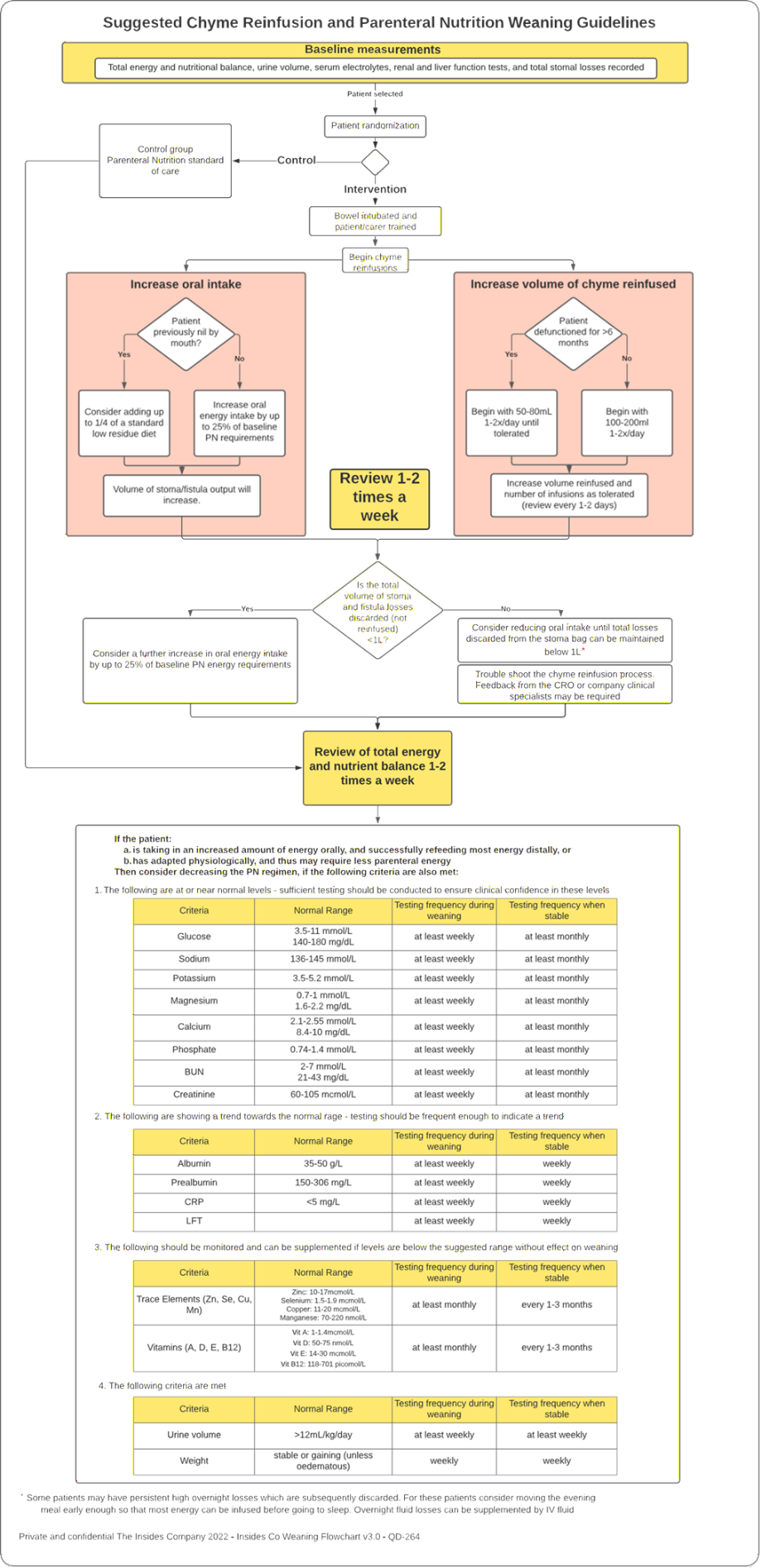

Guidelines for weaning parenteral nutrition

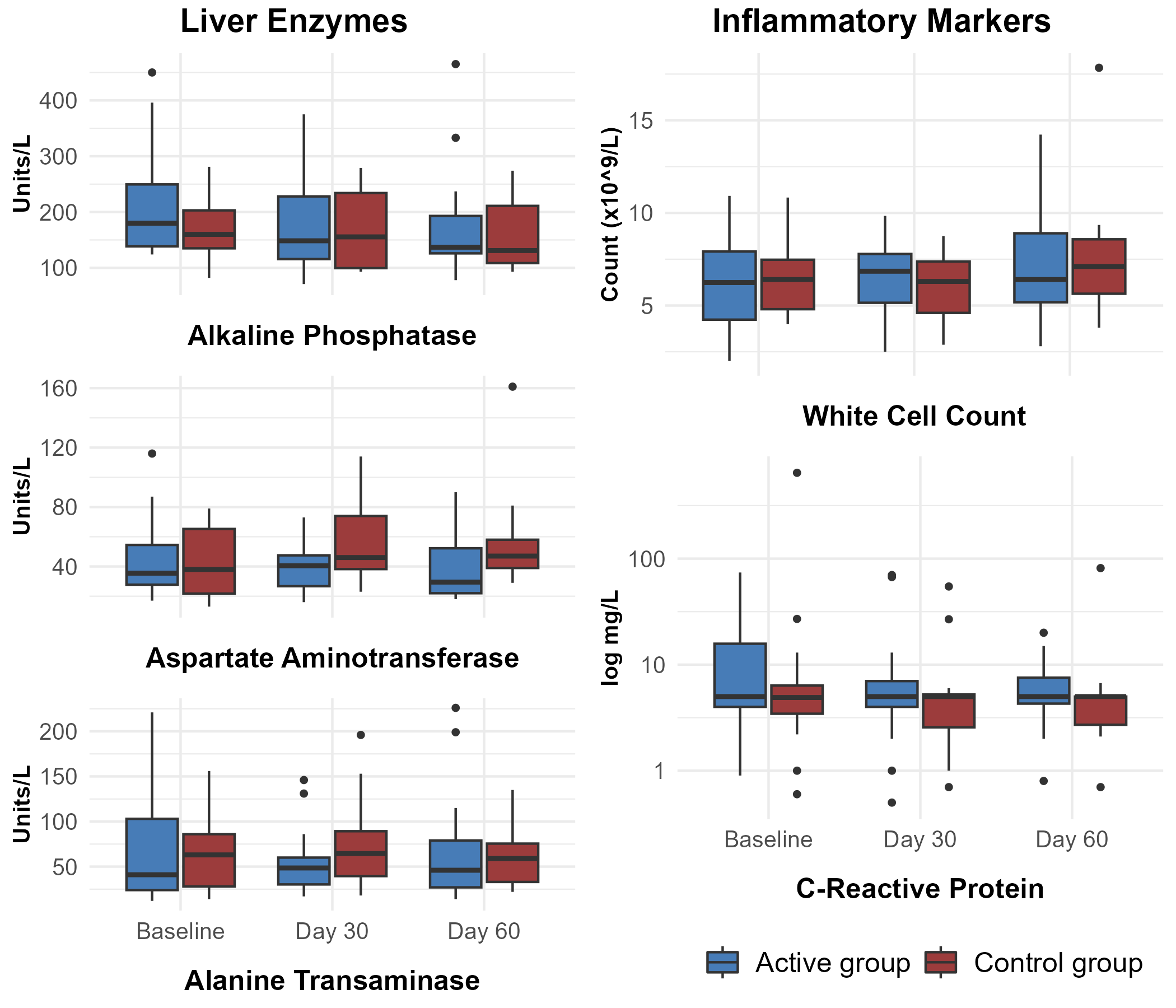
**Supplementary Figure 2**

Results of liver enzyme and inflammatory markers at baseline, Day 30, and Day 60. Result show median and interquartile range. There were no differences in the in-group and between-group comparisons (all p>0.05)

**Supplementary Table 2**

| **Site Name/Site #** | **No. Enrolled** | **No. Randomised** |
| --- | --- | --- |
| Henry Ford Health System (103) | 0 | 0 |
| University of Nebraska Medical Center (105) | 1 | 0 |
| University of Chicago (106) | 1 | 1 |
| University of Florida (107) | 1 | 0 |
| University of Miami (108) | 2 | 1 |
| St James's University Hospital, Leeds (201) | 13 | 11 |
| Royal Victoria Infirmary, Newcastle (202) | 2 | 2 |
| Nottingham City Hospital (203) | 8 | 8 |
| Southampton General Hospital (204) | 2 | 2 |
| St Mark's Hospital, London (205) | 3 | 3 |
| Royal Devon & Exeter Hospital (206) | 2 | 2 |
| Salford Royal Northern Care Alliance NHS Foundation Trust (207) | 9 | 9 |

**Supplementary Table 3**

| **Patient Satisfaction Questionnaire** | | |
| --- | --- | --- |
| Please read the following 10 questions carefully and score with one of five responses that range from Strongly Agree to Strongly Disagree. | | |
| 1. | I think that I would like to use this medical device frequently | 1 Strongly agree  2  3  4  5 Strongly disagree |
| 2. | I found the medical device unnecessarily complex | 1 Strongly agree  2  3  4  5 Strongly disagree |
| 3. | I thought the medical device was easy to use | 1 Strongly agree  2  3  4  5 Strongly disagree |
| 4. | I think that I would need the support of a technical person to be able to use this medical device | 1 Strongly agree  2  3  4  5 Strongly disagree |
| 5. | I found the various functions in this medical device were well integrated | 1 Strongly agree  2  3  4  5 Strongly disagree |
| 6. | I thought there was too much inconsistency in this medical device | 1 Strongly agree  2  3  4  5 Strongly disagree |
| 7. | I would imagine that most people would learn to use this medical device very quickly | 1 Strongly agree  2  3  4  5 Strongly disagree |
| 8. | I found the medical device very awkward to use | 1 Strongly agree  2  3  4  5 Strongly disagree |
| 9. | I felt very confident using the medical device | 1 Strongly agree  2  3  4  5 Strongly disagree |
| 10. | I needed to learn a lot of things before I could get going with this medical device | 1 Strongly agree  2  3  4  5 Strongly disagree |

Marking Schema

A device-positive response to the odd numbered statements appears to score 1 (Strongly agree) through to 5 (Strongly disagree), whereas a device-positive response to the even numbered statements applies the opposite score value. This was resolved by correcting the scores for all odd numbered statements to:

- 5 Strongly agree
- 4
- 3
- 2
- 1 Strongly disagree

No changes were made to the even numbered statement score values. This approach ensured the consistency of scoring such that the lower the individual statement scores, and the overall score, indicates a negative response to the Insides device. The higher the individual and overall scores, the latter to a maximum of 50, indicates a positive response to the Insides device.

**Supplementary Table 4**

| **Patient Satisfaction score through to Day 60**  **(ITT population Active Group N=26)** | | |
| --- | --- | --- |
| **Total score (10 - 50)**  **(Higher = more satisfied)** | **Active Group (N=26)** | |
|  | **Day 30** | **Day 60**  **or at closure if earlier** |
| **N (available)** | 23 | 21 |
| **Median (min, max)** | 48 (32, 50) | 46 (24, 50) |

**Supplementary Table 5**

| **Quality of Life Measures – Baseline to Day 60** | | | | |
| --- | --- | --- | --- | --- |
|  | **Active Group (N=26)** | | **Control Group (N=13)** | |
|  | **Observed** | **Change from Baseline** | **Observed** | **Change from Baseline** |
| **EQ-5D-3L Utility Score from Baseline to Day 60**  **(ITT population N=39)** | | | | |
| **Baseline** | | | | |
| **N** | 26 |  | 13 |  |
| **Mean (SD)** | 0.67 (0.25) |  | 0.64 (0.25) |  |
| **Day 30** | | | | |
| **N** | 23 | 23 | 12 | 12 |
| **Mean (SD)** | 0.72 (0.19) | 0.06 (0.23) | 0.73 (0.16) | 0.07 (0.23) |
| **Day 60 (or at closure if earlier)** | | | | |
| **N** | 21 | 21 | 10 | 10 |
| **Mean (SD)** | 0.69 (0.21) | -0.01 (0.26) | 0.74 (0.13) | 0.04 (0.21) |
| **Stoma QOL Scale from Baseline to Day 60  (ITT population N=39)** | | | | |
| **Baseline** | | | | |
| **N** | 26 |  | 13 |  |
| **Mean (SD)** | 49.8 (14.2) |  | 56.5 (12.2) |  |
| **Day 30** | | | | |
| **N** | 23 | 23 | 12 | 12 |
| **Mean (SD)** | 49.4 (13.8) | 0.2 (11.4) | 55.1 (13.4) | -2.3 (10.4) |
| **Day 60 (or at closure if earlier)** | | | | |
| **N** | 21 | 21 | 10 | 10 |
| **Mean (SD)** | 47.9 (13.7) | -3 (11.8) | 56 (11.9) | -5.5 (12.1) |
| **Beck Depression Inventory from Baseline to Day 60  (ITT population N=39)** | | | | |
| **Baseline** | | | | |
| **N** | 26 |  | 13 |  |
| **Mean (SD)** | 11.6 (8.4) |  | 11.7 (6.9) |  |
| **Day 30** | | | | |
| **N** | 23 | 23 | 12 | 12 |
| **Mean (SD)** | 12.3 (7.8) | 0.6 (6.8) | 10.6 (11.4) | -0.2 (7.7) |
| **Day 60 (or at closure if earlier)** | | | | |
| **N** | 21 | 21 | 10 | 10 |
| **Mean (SD)** | 11.4 (7.5) | 1.2 (4.6) | 9.7 (8.7) | 0.1 (6.8) |

**Supplementary Table 6**

| **Adverse Events - Consent to Day 60 (AT population N=40)** | | |
| --- | --- | --- |
|  | **Active Group (N=27)** | **Control Group (N=13)** |
| **Number of subjects with N(%)** | | |
| At least one AE | 19 (70.4) | 6 (46.2) |
| At least one SAE | 8 (29.6) | 1 (7.7) |
| At least one severe AE | 5 (18.5) | 1 (7.7) |
| At least one AE related to the device | 9 (33.3) | N/A |
| At least one SAE related to the device | 2 (7.4) | N/A |
| At least one unanticipated adverse device effect (UADE) | 0 (0.0) | N/A |
| Death | 1 (3.7) | 0 (0.0) |
| **Number of events N** | | |
| Number of AEs reported | 35 | 15 |
| Number of SAEs reported | 14 | 2 |
| Number of severe AEs reported | 7 | 2 |
| Number of AEs related to the device | 11 | N/A |
| Number of SAEs related to the device | 2 | N/A |

**Supplementary Table 7**

| **Adverse Event types unrelated to Device - Consent to Day 60**  **(AT population N=40)** | | | | | | |
| --- | --- | --- | --- | --- | --- | --- |
| **MedDRA Preferred Term** | **Active Group (N=27)** | | **Control Group (N=13)** | | **Total (N=40)** | |
|  | **Events N** | **Subjects N(%)** | **Events N** | **Subjects N(%)** | **Events N** | **Subjects N(%)** |
| Abdominal pain | 1 | 1 (3.7) | 2 | 2 (15.4) | 3 | 3 (7.5) |
| Acute kidney injury | 1 | 1 (3.7) | 2 | 2 (15.4) | 3 | 3 (7.5) |
| Alkaline phosphatase increased | 1 | 1 (3.7) | 0 | 0 (0.0) | 1 | 1 (2.5) |
| Bacteremia | 1 | 1 (3.7) | 0 | 0 (0.0) | 1 | 1 (2.5) |
| Dehydration | 2 | 2 (7.4) | 0 | 0 (0.0) | 2 | 2 (5.0) |
| Diabetes mellitus | 0 | 0 (0.0) | 1 | 1 (7.7) | 1 | 1 (2.5) |
| Headache | 1 | 1 (3.7) | 0 | 0 (0.0) | 1 | 1 (2.5) |
| Hepatobiliary disorders - Other, specify | 1 | 1 (3.7) | 1 | 1 (7.7) | 2 | 2 (5.0) |
| Hyperphosphatemia | 1 | 1 (3.7) | 0 | 0 (0.0) | 1 | 1 (2.5) |
| Hypokalemia | 1 | 1 (3.7) | 0 | 0 (0.0) | 1 | 1 (2.5) |
| Injury, poisoning and procedural complications - Other, specify | 1 | 1 (3.7) | 0 | 0 (0.0) | 1 | 1 (2.5) |
| Intestinal stoma leak | 0 | 0 (0.0) | 1 | 1 (7.7) | 1 | 1 (2.5) |
| Metabolism and nutrition disorders - Other, specify | 0 | 0 (0.0) | 1 | 1 (7.7) | 1 | 1 (2.5) |
| Myelitis | 1 | 1 (3.7) | 0 | 0 (0.0) | 1 | 1 (2.5) |
| Nausea | 0 | 0 (0.0) | 1 | 1 (7.7) | 1 | 1 (2.5) |
| Neoplasms benign, malignant and unspecified (incl cysts and polyps) - Other, specify | 0 | 0 (0.0) | 1 | 1 (7.7) | 1 | 1 (2.5) |
| Otitis externa | 1 | 1 (3.7) | 0 | 0 (0.0) | 1 | 1 (2.5) |
| Pneumonia | 1 | 1 (3.7) | 0 | 0 (0.0) | 1 | 1 (2.5) |
| Prolapse of intestinal stoma | 1 | 1 (3.7) | 0 | 0 (0.0) | 1 | 1 (2.5) |
| Sepsis | 3 | 3 (11.1) | 0 | 0 (0.0) | 3 | 3 (7.5) |
| Skin and subcutaneous tissue disorders - Other, specify | 1 | 1 (3.7) | 1 | 1 (7.7) | 2 | 2 (5.0) |
| Small intestinal obstruction | 0 | 0 (0.0) | 1 | 1 (7.7) | 1 | 1 (2.5) |
| Vascular access complication | 1 | 1 (3.7) | 1 | 1 (7.7) | 2 | 2 (5.0) |
| Vascular disorders - Other, specify | 2 | 2 (7.4) | 1 | 1 (7.7) | 3 | 3 (7.5) |
| Vomiting | 0 | 0 (0.0) | 1 | 1 (7.7) | 1 | 1 (2.5) |
| Wound complication | 2 | 2 (7.4) | 0 | 0 (0.0) | 2 | 2 (5.0) |
| **Event Severity** | | | | | | |
| Mild | 9 | 5 (18.5) | 9 | 5 (38.5) | 18 | 10 (25) |
| Moderate | 9 | 7 (25.9) | 4 | 1 (7.7) | 13 | 8 (20) |
| Severe | 6 | 4 (14.8) | 2 | 1 (7.7) | 8 | 5 (12.5) |
| **Frequency** | | | | | | |
| Single episode | 16 | 9 (33.3) | 11 | 6 (46.2) | 27 | 15 (37.5) |
| Intermittent | 4 | 4 (14.8) | 1 | 1 (7.7) | 5 | 5 (12.5) |
| Continuous | 4 | 3 (11.1) | 3 | 3 (23.1) | 7 | 6 (15) |
| **Relationship to procedure** | | | | | | |
| Definitely related | 1 | 1 (3.7) | 3 | 2 (15.4) | 4 | 3 (7.5) |
| Possibly related | 0 | 0 (0.0) | 0 | 0 (0) | 0 | 0 (0) |
| Not related | 23 | 13 (48.1) | 12 | 6 (46.2) | 35 | 19 (47.5) |
| **Action Taken** | | | | | | |
| Required medication | 12 | 8 (29.6) | 1 | 1 (7.7) | 13 | 9 (22.5) |
| Surgical Intervention | 3 | 3 (11.1) | 1 | 1 (7.7) | 4 | 4 (10) |
| Other | 12 | 8 (29.6) | 11 | 6 (46.2) | 23 | 14 (35) |
| None | 1 | 1 (3.7) | 6 | 3 (23.1) | 7 | 4 (10) |
| **Outcome** | | | | | | |
| Resolved | 18 | 10 (37.0) | 11 | 5 (38.5) | 29 | 15 (37.5) |
| Resolved with sequelae | 3 | 3 (11.1) | 0 | 0 (0.0) | 3 | 3 (7.5) |
| Ongoing | 2 | 2 (7.4) | 4 | 3 (23.1) | 6 | 5 (12.5) |
| Death | 1 | 1 (3.7) | 0 | 0 (0) | 1 | 1 (2.5) |

**Supplementary Table 8**

| **Device Related Adverse Event types - Consent to Day 60**  **(AT population Active Group N=27)** | | |
| --- | --- | --- |
| **MedDRA Preferred Term** | **As Treated Active Group (N=27)** | |
|  | **Events (N)** | **Subjects N (%)** |
| Abdominal pain | 2 | 2 (7.4) |
| General disorders and administration site conditions - Other, specify | 5 | 3 (11.1) |
| Injury, poisoning and procedural complications - Other, specify | 1 | 1 (3.7) |
| Prolapse of intestinal stoma | 1 | 1 (3.7) |
| Skin and subcutaneous tissue disorders - Other, specify | 1 | 1 (3.7) |
| Skin ulceration | 1 | 1 (3.7) |
| **Event Severity** | | |
| Mild | 6 | 4 (14.8) |
| Moderate | 4 | 4 (14.8) |
| Severe | 1 | 1 (3.7) |
| **Frequency** | | |
| Single episode | 4 | 3 (11.1) |
| Intermittent | 3 | 3 (11.1) |
| Continuous | 4 | 4 (14.8) |
| **Relationship to procedure** | | |
| Definitely related | 1 | 2 (3.7) |
| Possibly related | 2 | 2 (7.4) |
| Not related | 7 | 7 (25.9) |
| Unknown | 1 | 1 (3.7) |
| **Relationship to device** | | |
| Definitely related | 6 | 5 (18.5) |
| Possibly related | 5 | 5 (18.5) |
| **Action Taken** | | |
| Required medication | 1 | 1 (3.7) |
| Surgical Intervention | 0 | 0 (0.0) |
| Other | 11 | 9 (33.3) |
| **Outcome** | | |
| Resolved | 9 | 7 (25.9) |
| Resolved with sequelae | 2 | 2 (7.4) |
